# Latin-American Registry of Cardiovascular Disease and COVID-19: Final Results

**DOI:** 10.1101/2023.03.15.23287304

**Authors:** Juan Esteban Gómez-Mesa, Stephania Galindo, Manuela Escalante-Forero, Yorlany Rodas, Andrea Valencia, Eduardo Perna, Alexander Romero, Iván Mendoza, Fernando Wyss, José Luis Barisani, Mario Speranza, Walter Alarco, Noel Alberto Flórez

## Abstract

**Background:** COVID-19 is a global disease caused by the Severe Acute Respiratory Syndrome Coronavirus 2 (SARS-CoV-2). Patients with a severe or critical illness can develop respiratory and cardiovascular complications. This study aimed to describe a Latin American and Caribbean (LA&C) population with COVID-19 to provide information related to this disease, in-hospital cardiovascular complications and in-hospital mortality.

**Methods:** The CARDIO COVID-19-20 Registry is an observational, multicenter, ambispective, and hospital-based registry of patients with confirmed COVID-19 infection that required in-hospital treatment in LAC. Enrollment of patients started on May 01, 2020, and ended on June 30, 2021.

**Results:** The CARDIO COVID-19-20 Registry included 3260 patients from 44 institutions of 14 LA&C countries. 63.2% patients were male and median age was 61.0 years old. Most common comorbidities were overweight/obesity (49.7%), hypertension (49.0%), and diabetes mellitus (26.7%). Most frequent cardiovascular complications were cardiac arrhythmia (9.1%), decompensated heart failure (8.5%), and pulmonary embolism (3.9%). 53.5% of patients were admitted to Intensive Care Unit (ICU), and median length of stay at the ICU was 10.0 days. Support required in ICU included invasive mechanical ventilation (34.2%), vasopressors (27.6%), inotropics (10.3%) and vasodilators (3.7%). Rehospitalization after 30-day post discharge was 7.3%. In-hospital mortality and 30-day post discharge was 25.5% and 2.6%, respectively.

**Conclusions:** The LA&C population with COVID-19 patients and hospitalization, has a considerable burden of cardiovascular diseases related to a worse prognosis. It is necessary to carry out a more specific analysis to determine risk factors for cardiovascular outcome.

## Background

The Severe Acute Respiratory Syndrome Coronavirus 2 (SARS-CoV-2) can spread through respiratory droplets (it can stay contagious and can be suspended in the air for up to three hours), human-to-human transmission, by contaminated objects, and airborne contagion. The reported contagion rates from a patient with symptomatic infection vary by location and efficiency of infection control measures (1). The current gold standard for diagnosing COVID-19 requires polymerase chain reaction (PCR) for the detection of the viral genetic material (RNA) in a nasopharyngeal swab or sputum sample (2). The serological testing for the-CoV-2 immunoglobulin G (IgG) and/or IgM is widely used in the diagnosis of COVID-19, a meta-analysis showed that the detection of anti-SARS-CoV-2 IgG and IgM had high diagnostic efficiency to assist the diagnosis of SARS-CoV-2 (3). In patients with suspected COVID-19, the diagnosis cannot rely only on PCR test results, it also has to rely on clinical presentation and the findings from other tests, most notably chest computed tomography (CT) (4).

The coronavirus disease 2019 (COVID-19) pandemic is caused by SARS-CoV-2, an RNA virus that turned into a major public health concern after the outbreak of the Middle East Respiratory Syndrome-CoV (MERS-CoV) and severe acute respiratory syndrome-CoV (SARS-CoV) in 2002 and 2012, respectively. In total, seven human coronaviruses (HCoVs) have now been discovered, but the definitive origin of SARS-CoV-2 remains undetermined (5,6).

The COVID-19 pandemic started in Wuhan, China in late 2019; the epicenter moved across the world, then arrived at Latin America and Caribbean (LA&C) where the first case of COVID-19 was confirmed on February 25, 2020. According to the World Health Organization (7), until February of 2023 more than 753 million cases have been reported worldwide, 188 million of them from the Americas. Some of the most affected LA&C countries are Brazil (36.7 million), Argentina (10.0 million), Mexico (7.3 million), and Colombia (6.3 million). Socioeconomic factors contribute to a more severe impact of COVID-19 in this region than in developed countries; moreover, some of the comorbidities associated with a worse prognosis of COVID-19, such as obesity, diabetes, and hypertension are highly prevalent in LA&C (8–13).

Based on the above, the CARDIOCOVID-19-20 Registry collected information of hospitalized patients with COVID-19 from 14 LA&C countries, this article aimed to describe demographics, general information, and in-hospital findings of this population. Other specific analysis will be developed in further publications.

## Methods and Design

The CARDIO COVID-19-20 Registry is an observational, multicenter, ambispective, and hospital-based registry that included patients with confirmed COVID-19 that required in-hospital treatment in LA&C institutions (14). Enrollment of patients started on May 2020 and ended in June 2021. The Biomedical Research Ethics Committee of the Fundación Valle del Lili in Cali, Colombia approved this study under act number 409 – 2021. Informed consent was not required because of the observational design. A 30-day follow-up after hospital discharge was planned for every recruited patient.

The following countries participated in the CARDIO COVID-19-20 Registry (in alphabetical order): Argentina, Brazil, Chile, Colombia, Costa Rica, Ecuador, El Salvador, Guatemala, Mexico, Panama, Paraguay, Peru, Dominican Republic, and Venezuela.

Criteria to participate in CARDIO COVID-19-20 Registry included those patients older than 18 years with a confirmed diagnosis of COVID-19 according to guidelines given by the World Health Organization and according to each institutional and/or local guideline, who required in-hospital management for more than 24 hours, or who died during the first 24 hours after hospital admission.

The data was collected in the electronic database system REDCap (Research Electronic Data Capture: https://www.project-redcap.org/). This database system included 277 variables. The confidentiality of patient data was assured, each institution was coded, as well as the identity of every enrolled patient.

### Data Analysis

Demographics and clinical characterization of the in-hospital population was carried out through a univariate analysis. Variables with normal distribution were identified with a Shapiro Wilk test. Categorical variables are reported as a number and percentage, normally distributed continuous variables as mean with Standard Deviation (SD), and variables with skewed distribution as median with Interquartile Range (IQR). Clinical variables analyzed included paraclinics (at admission and at discharge), pathological history, cardiovascular treatment prior to admission, clinical manifestations at admission, clinical findings at admission, cardiovascular complications during hospitalization, cardiac imaging tests, cardiovascular procedures performed during hospitalization, medication used for COVID-19, ICU admission, in-hospital deaths and post-discharge 30-day deaths. All analyses were performed using the Stata 17.0 MP version.

## Results

The CARDIO COVID-19-20 Registry included 3404 patients in the database, but 144 patients were excluded from the final analysis due to unresolved queries and/or data inconsistencies, resulting in 3260 patients from 44 institutions located in 14 Caribbean, Central and South American countries (Argentina, Brazil, Chile, Colombia, Costa Rica, Ecuador, El Salvador, Guatemala, Mexico, Panama, Paraguay, Peru, Dominican Republic, and Venezuela) (Figure 1). 2059 were male (63.2%) and median age was 61.0 years old. All included patients had a confirmed diagnosis of COVID-19 and 120 (3.8%) of them were healthcare workers (Table 1). Exposure type was classified as by circulation (58.8), followed by contact (39.3%) and lastly imported (1.9%). We defined the type of infection as “circulation” when the patient did not know where the infection was acquired, as “contact” when the patient was in close contact with someone with COVID-19 and knew who infected them, and as “imported” when the patient acquired the infection in a foreign country and then traveled back to his/her country.

**Table 1.**
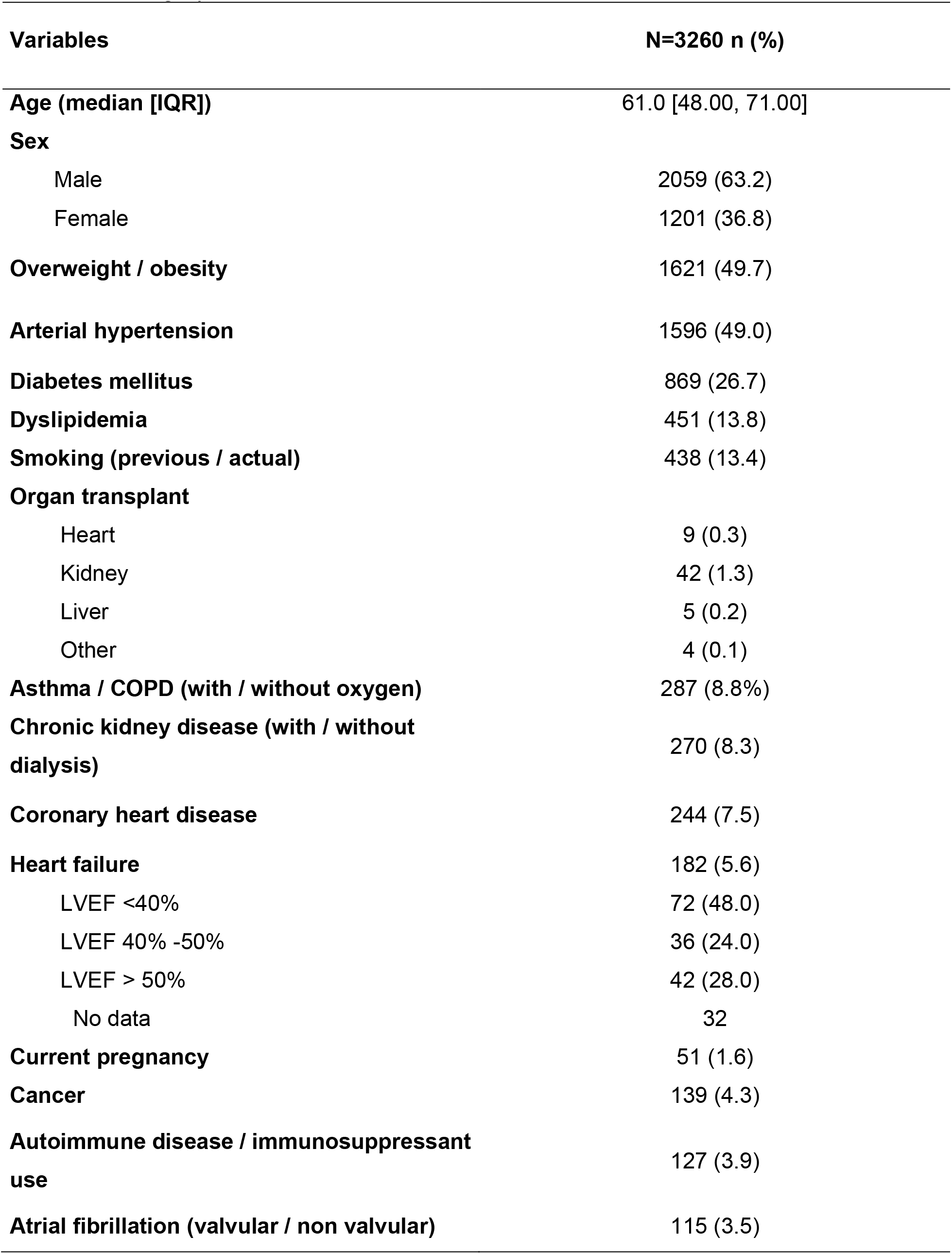
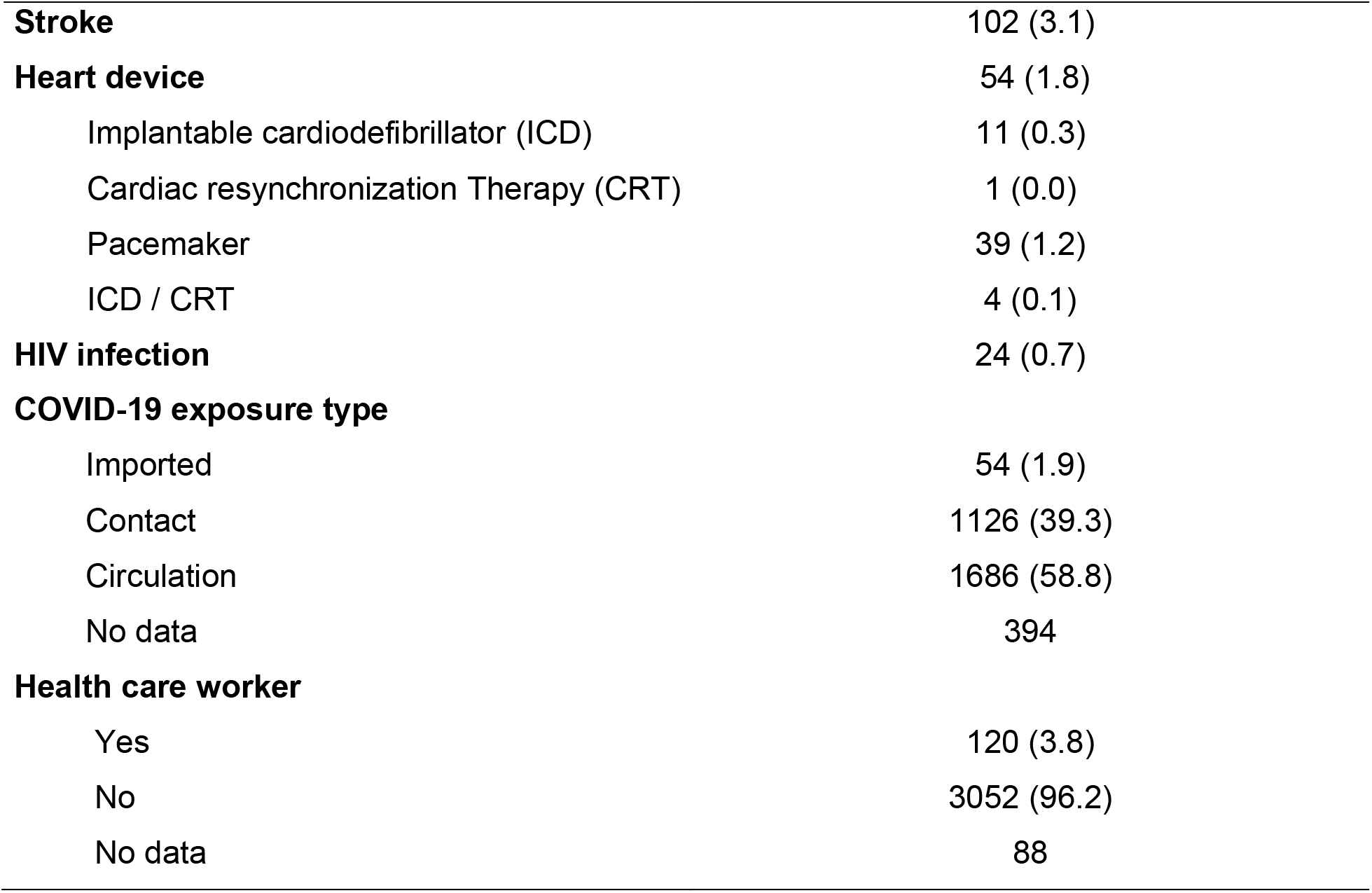
Demographic and comorbidities

**Figure 1.**
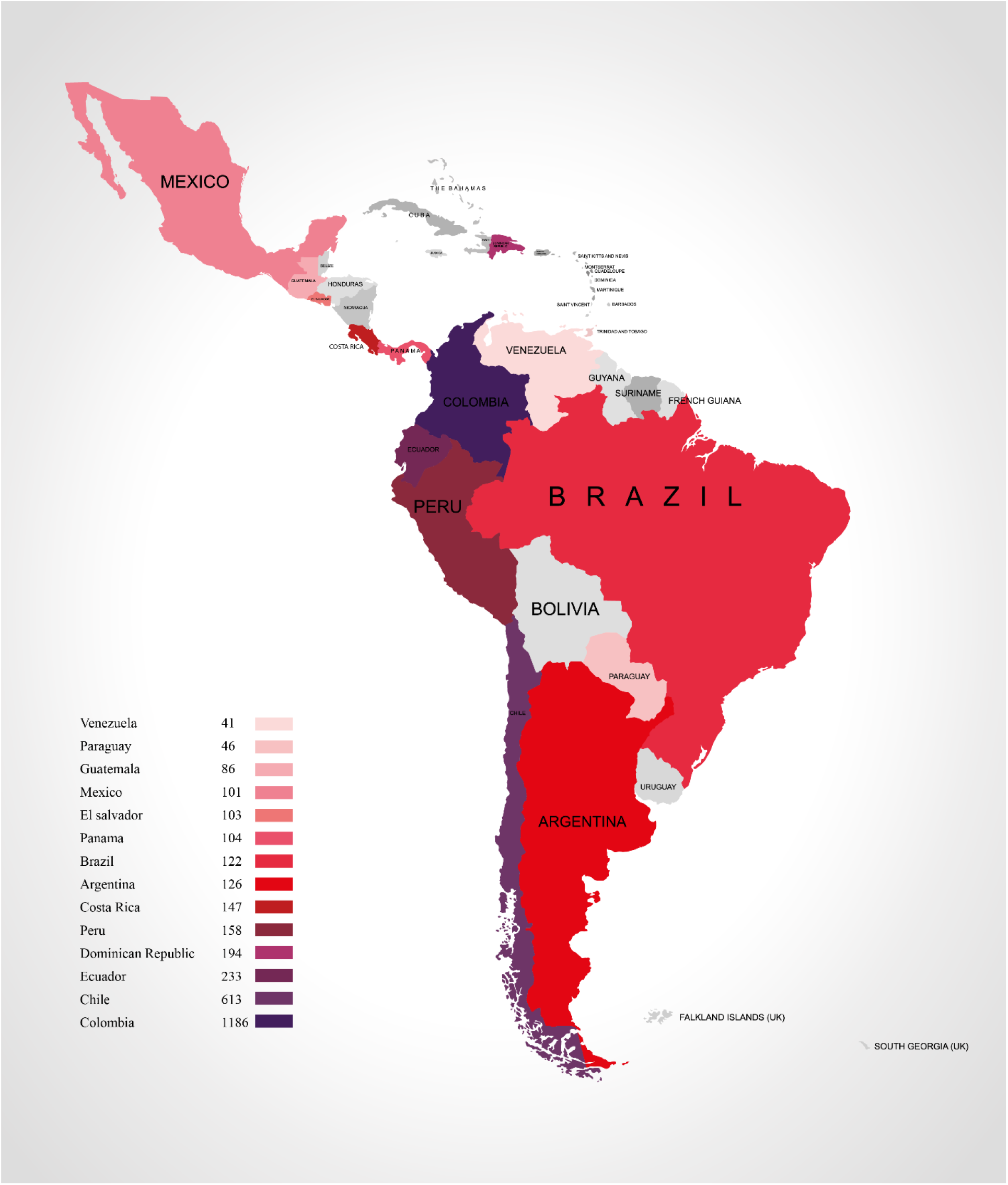
Number of patients by participant countries

The most common preexisting comorbidities were overweight/obesity (49.7%), hypertension (49.0%), diabetes mellitus (26.7%) and dyslipidemia (13.8%) (Table 1). 12-lead electrocardiogram was performed in 1626 patients (49.9%), and abnormal findings included right bundle branch block (6.5%), atrial fibrillation (5.4%), left bundle branch block (3.1%) and ventricular extrasystoles (1.7%) (Table 2).

**Table 2.** Diagnostic tests

| **Variables** | **n (%)** |
| --- | --- |
| **ECG performed** | 1626 (49.9) |
| **Rhythm, n=1556** |  |
| Sinus rhythm | 1377 (88.5) |
| Ventricular extrasystole | 27 (1.7) |
| Supraventricular extrasystole | 9 (0.6) |
| Atrial fibrillation | 84 (5.4) |
| Atrial flutter | 7 (0.4) |
| Other | 52 (3.3) |
| **QRS complex (median [IQR]), n=1,393** | 90.00 [80.00, 102.00] |
| **Bundle Branch Block, n= 1,515** |  |
| Right | 98 (6.5) |
| Left | 47 (3.1) |
| **Corrected QTc interval (median [IQR]), n=1,386** | 419.0 [395.0, 443.0] |
| **Echocardiogram performed** | 634 (19.4) |
| **Systolic function, n=611** |  |
| Focally decreased | 51 (8.3) |
| Globally decreased | 96 (15.7) |
| Normal | 444 (72.7) |
| **Right ventricular dysfunction, n= 589** | 96 (16.3) |
| **Pericardial effusion, n= 625** |  |
| Mild | 57 (9.1) |
| Moderate | 9 (1.4) |
| Severe | 2 (0.3) |
| **Cardiac tamponade, n=618** | 2 (0.3) |
| **Severe valvular insufficiency, n=609** |  |
| Mitral | 11 (1.8) |
| Aortic | 3 (0.5) |
| Both | 2 (0.3) |
| **Severe valvular stenosis, n=610** |  |
| Mitral | 5 (0.8) |
| Aortic | 12 (2.0) |
| Both | 0 (0.0) |
| **Inferior vena cava dilatation, n=576** | 90 (15.6) |
| **LVEF (median [IQR]), n=621** | 60.00 [50.00, 65.00] |
| **TAPSE (median [IQR]), n=365** | 20.00 [17.00, 23.00] |
| **PSAP (median [IQR]), n=357** | 35.00 [28.00, 45.00] |
| **Strain longitudinal (median [IQR]), n=33** | 18.00 [16.00, 21.00] |
| **Diameter diastolic vent left (median [IQR]), n=350** | 46.00 [42.00, 52.00] |
| **Systolic diameter vent left (median [IQR]), n=322** | 30.00 [27.00, 37.00] |
| **Chest X-ray performed** | 3019 (92.6) |
| **Pulmonary infiltrates** |  |
| Unilateral | 239 (7.3) |
| Bilateral | 2366 (72.6) |
| **Cardiomegaly** | 549 (16.8) |
| **Pulmonary congestion** |  |
| Unilateral | 75 (2.3) |
| Bilateral | 533 (16.3) |
| **Pleural effusion** |  |
| Unilateral | 170 (5.2) |
| Bilateral | 190 (5.8) |

Chest x-ray was performed in 3019 patients (92.6%), and abnormal findings included pulmonary infiltrates (79.9%), pulmonary congestion (18.7%), cardiomegaly (16.8%) and pleural effusion (11.0%) (Table 2). Echocardiogram was performed in 634 patients (19.4%), and abnormal findings included globally decreased systolic function (15.7%), right ventricular dysfunction (16.3%), pericardial effusion (10.9%) and inferior vena cava dilatation (15.6%) (Table 2).

Paraclinical tests at admission showed median leukocytes count of 8710.0mm^3^ [6310.0, 12 125.0], median hemoglobin value of 13.6gr/dl [12.1, 14.9], median platelets count of 228 500.0µl [176 000.0, 297 000.0], median creatinine levels of 0.9mg/dl [0.7, 1.3], and median NT-proBNP level of 429.0pg/ml [88.0, 2744.5] (Supplementary material). In contrast, paraclinical tests at discharge showed differences in median lymphocytes level (1319.5mm^3^ [800.0, 1970.0]), median hemoglobin value (12.1gr/dl [10.3, 13.7]), median hematocrits value (36.5%), and median platelets count (283 000.0µl [202 000.0, 378 000.0]) (Supplementary material).

The most common cardiovascular complications during hospitalization were cardiac arrhythmia (9.1%), decompensated heart failure (8.5%), pulmonary embolism (3.9%), and acute coronary syndrome (2.9%). 53.5% of patients were admitted to the ICU and median length of stay at the ICU was 10.0 days [5.0, 18.0]. Invasive mechanical ventilation (IMV) was required in 34.2% of patients, while 15.1% required non-invasive MV (NiMV); vasopressors were needed in 27.6% of patients, inotropics in 10.3% of patients and vasodilators in 3.7% of patients. Other procedures performed during hospitalization included central venous catheter (20.0%), cardioversion / defibrillation (3.3%), coronary arteriography (1.6%), coronary fibrinolysis / thrombolysis (1.0%), coronary angioplasty (1.0%), use of extracorporeal oxygenation membrane (ECMO) (0.9%) and intraaortic balloon pump (0.1%) (Table 3).

**Table 3.** In-hospital interventions

| **Variable** | **N=3260 n (%)** |
| --- | --- |
| **Invasive Mechanical Ventilation (IMV)** | 1115 (34.2) |
| IMV days (median [IQR]) | 11.00 [6.00, 19.00] |
| **Vasopressor use** | 900 (27.6) |
| Vasopressor use days (median [IQR]) | 7.00 [3.00, 12.80] |
| **Inotropic use** | 336 (10.3) |
| Inotropic use days (median [IQR]) | 5.00 [3.00, 10.00] |
| **Vasodilator use** | 119 (3.7) |
| Vasodilator use days (median [IQR]) | 3.00 [2.00, 6.00] |
| **Central venous catheter** | 651 (20.0) |
| Central venous catheter days (median [IQR]) | 12.00 [6.00, 20.00] |
| **Cardioversion / defibrillation** | 106 (3.3) |
| **Coronary arteriography** | 52 (1.6) |
| **Coronary fibrinolysis / thrombolysis** | 31 (1.0) |
| **Coronary angioplasty** | 31 (1.0) |
| **Extracorporeal Oxygenation Membrane (ECMO)** | 29 (0.9) |
| ECMO days (median [IQR]) | 17.00 [8.00, 28.00] |
| **Intraaortic balloon pump (IABP)** | 2 (0.1) |
| IABP days (median [IQR]) | 5.00 [4.50, 5.50] |
| **Intensive Care Unit (ICU)** | 1745 (53.5) |
| ICU days (median [IQR]) | 10.00 [5.00, 18.00] |

The most common pharmacological treatment for COVID-19 patients included corticosteroids (67.4%), thromboprophylaxis (62.0%), anticoagulation therapy (38.6%), azithromycin (33.6%) and hydroxychloroquine (21.2%) (Table 4).

**Table 4.** Pharmacological treatment for COVID-19 during hospitalization

| **Medicaments\*** | **N=3260 n (%)** |
| --- | --- |
| **Corticosteroids** |  |
| Oral | 154 (4.7) |
| Parenteral | 2043 (62.7) |
| **Thromboprophylaxis** | 2021 (62.0) |
| **Anticoagulation** | 1257 (38.6) |
| **Azithromycin** | 1096 (33.6) |
| **Hydroxychloroquine** | 690 (21.2) |
| **Lopinovir** | 234 (7.2) |
| **Ritonavir** | 229 (7.0) |
| **Chloroquine** | 90 (2.8) |
| **Plasmapheresis** | 31 (1.0) |
| **Immunoglobulin** | 23 (0.7) |
| **Interferon** | 9 (0.3) |
| **Other** | 1189 (36.5) |

\*More than one medicament can be administered to patients.

In-hospital mortality was 25.5%, being mainly non-cardiovascular, and 30-day rehospitalization and mortality after discharge was 7.3% and 2.6%, respectively (Table 5).

**Table 5.**
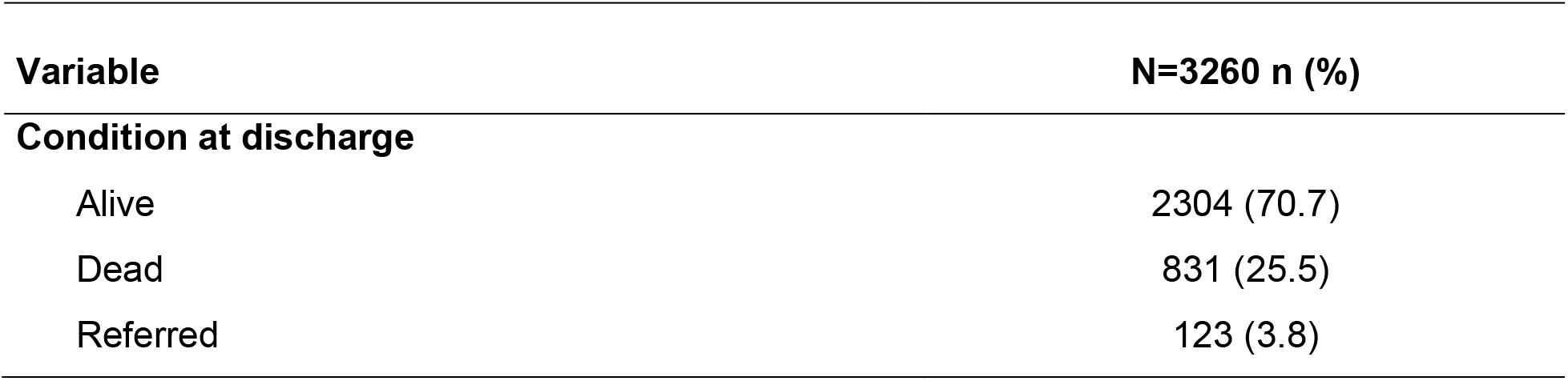
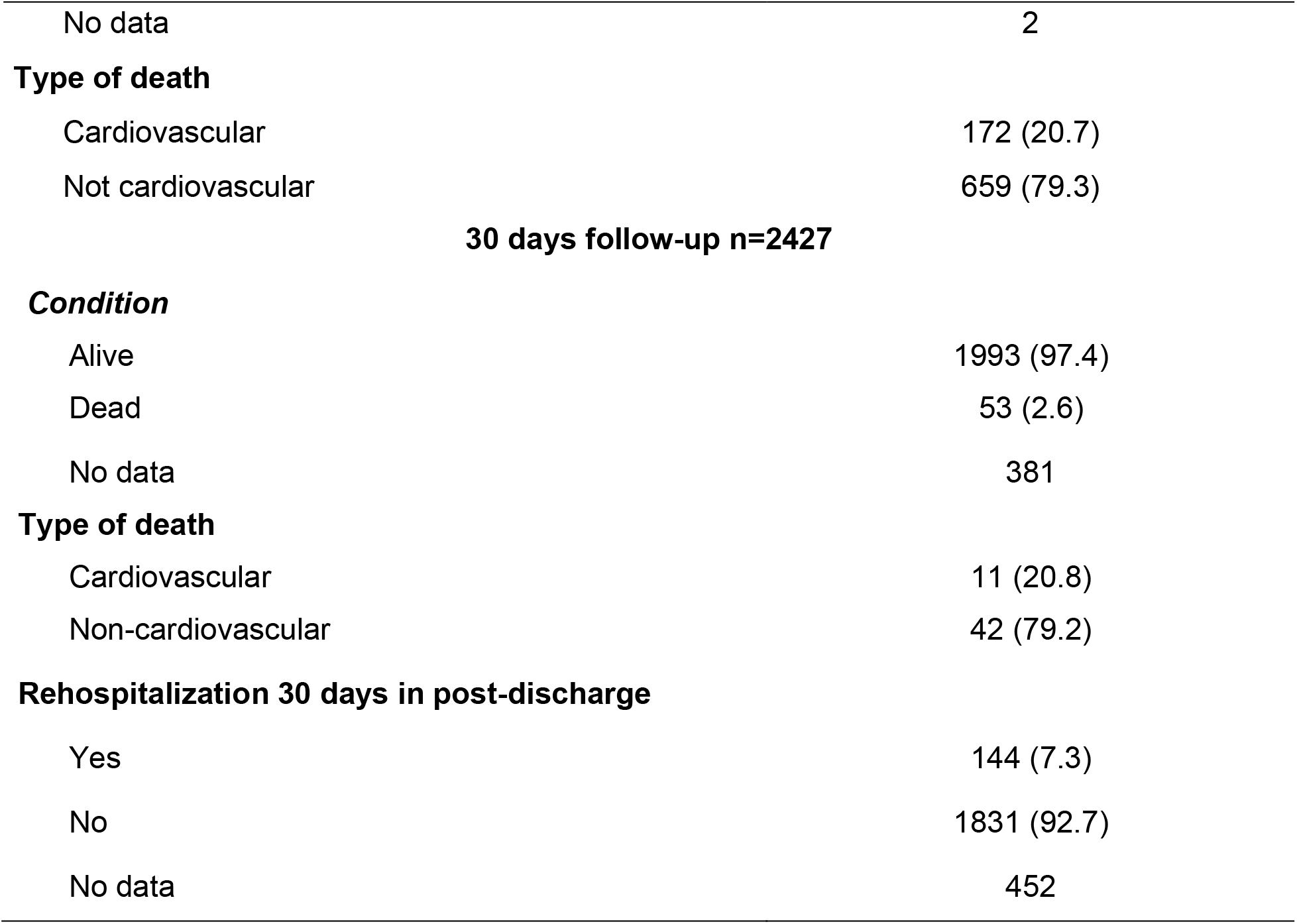
Outcomes during hospitalization and 30-day follow up

## Discussion

There are still many unsolved questions about COVID-19 and reported data until today can vary from one population to another. To be able to compare different populations and draw conclusions, each research group must share their knowledge and findings.

The SEMI-COVID-19 Registry provide data from more than 25 000 patients with COVID-19 hospitalized in Spain; 150 hospitals were included in this registry, mean age was 69.4 years. The most common comorbidities were hypertension (50.9%), dyslipidemia (39.7%), smoking history (30.7%), obesity (21.2%), and diabetes mellitus (19.4%) (9). Similar information was reported in our population regarding hypertension; however, overweight/obesity and diabetes mellitus percentages were significantly higher in our study, and dyslipidemia and smoking history were lower in our study. Obesity is related to a subclinical inflammatory state caused by an excess of inflammatory cells in adipose tissue. A meta-analysis found a 37.6% prevalence of pre-existing obesity in COVID-19 patients with poor outcomes, also, a higher mortality rate in obese patients who had other comorbidities, the most prevalent being cardiovascular disease, hypertension, and respiratory diseases (10). It has been described that COVID-19-related mortality in people with DM was higher in males and directly associated with age, cardiovascular, and renal complications, as well as with poorer glycemic control and higher BMI (11).

The study by Bilaloglu et al. (12) reported data from 3334 COVID-19 patients in New York who presented a median age of 64 years and 39.6% were female, similar information was found in our study. This population reported greater thrombotic events, 16% of patients had any thrombotic event, where 3.2% had pulmonary embolism (PE), 3.9% had deep vein thrombosis (DVT) and 11.1% had arterial events such as myocardial infarction, ischemic stroke, and other systemic thrombi (12).

The risk of developing venous thromboembolic events in critically ill patients is higher in the presence of COVID-19; the increased risk of thromboembolic events is also related to hemostatic alterations, immobility, systemic inflammatory status, mechanical ventilation, and central catheters. Several studies support the increased incidence of venous thromboembolic events in COVID-19 patients and their risk factors (13). 62.0% of our patients received thromboprophylaxis; according to evidence, thrombosis rates are lower in patients who received pharmacologic thromboprophylaxis (15).

Another cohort reported that 17% of COVID-19 patients required IMV, 14% non-IMV, and ECMO 2% (16); in contrast with our study where 34.2% of patients required IMV, and 0.9% of patients ECMO. These differences could be attributable to multiple variables, and the risk factors for these therapies must be reviewed.

A severe illness is characterized by diffuse alveolar damage, inflammatory infiltrates, and microvascular thrombosis. COVID-19 is associated with diffuse lung damage, and glucocorticoids may modulate inflammation-mediated lung injury and thereby reduce progression to respiratory failure and death; 67.4% of patients from our study received treatment with corticosteroids. A randomized study showed significantly lower mortality at 28 days with dexamethasone treatment (17,18). It has been described to have the largest benefit among patients with IMV (18).

Stroke is another complication described among COVID-19 patients, during a retrospective study with 844 patients at three Philadelphia hospitals, authors found that 2.4% patients had ischemic stroke and 0.9% had intracranial hemorrhage. Most patients with ischemic stroke had conventional vascular risk factors, and traditional stroke mechanisms were common (19). Due to pro-inflammatory and hypercoagulation state during COVID-19 infection, the risk of stroke is higher, and it is expected to worsen outcomes of patients having pre-existing cerebrovascular diseases (20).

The SEMI-COVID-19 registry reported a lower number of deaths related to COVID-19 than our registry (21% vs 25.5%). We also had a greater number of rehospitalization than reported in SEMI-COVID-19 (3.9% vs 7.3%).

## Conclusions

We report in this LA&C population of hospitalized patients with COVID-19 a high burden of cardiovascular comorbidities related to a worse cardiovascular prognosis. More than half of these population required management in intensive care, with higher requirement of invasive mechanical ventilation and vasoactive support, resulting in a high in-hospital mortality and a considerable high 30-day post discharge rehospitalization and mortality.

More detailed analysis between comorbidities, in-hospital cardiovascular outcomes, mortality and rehospitalization will be done in order to implement future local and regional interventions for a better approach and treatment of these population.

## Data Availability

The authors confirm that the data supporting the findings of this study are available within the article and its supplementary materials.

## Ethics and consent

The Biomedical Research Ethics Committee of the Fundación Valle del Lili approved this study under act number 409 – 2021. Informed consent was not required because of the observational design. If any institution required informed consent, a form validated by the CIC and the Ethics Committee of the Fundación Valle del Lili was available.

## Funding Information

None

## Competing Interests

The authors have no competing interest to declare.

## Author contributions

JEG contributed to the conceptualization, data curation, formal analysis, investigation, methodology, project administration, supervision, validation, writing and editing the manuscript. SG contributed to the data curation, formal analysis, investigation, project administration, supervision, writing and editing the manuscript. ME contributed to the data curation, formal analysis, project administration, writing and editing the manuscript. YR contributed to the data curation, formal analysis, investigation, methodology, supervision, writing and editing the manuscript. EP, AR, IM, FW, JLB, MS, WA, and NAF contributed to the investigation. All authors read and approved the final manuscript.

## Notes

### Competing Interest Statement

The authors have declared no competing interest.

### Funding Statement

This study did not receive any funding

### Author Declarations

Ethics committee/IRB of Fundación Valle del Lili gave ethical approval for this work

