## Supplementary material for "Latin-American Registry of Cardiovascular Disease and COVID-19: Final Results"

Supplementary material. Paraclinic test results

| | Paraclinic test results | | | |
|---|---|---|---|---|
| **Variable** | **Paraclinics at admission n (%)** | **N** | **Paraclinics at discharge n (%)** | **N** |
| Complete blood count | | 3260 | | 3260 |
| 0. No | 22 (0.7) | | 365 (11.2) | |
| 1. Yes | 3238 (99.3) | | 2895 (88.8) | |
| Leukocytes_mm3 (median[IQR]) | 8710.0 (6310.0, 12125.0) | 3235 | 8640.0 (6207.5, 12500.0) | 2888 |
| Lymphocytes_mm3 (median[IQR]) | 1030.0 (700.0, 1480.0) | 3166 | 1319.5 (800.0, 1970.0) | 2838 |
| Hemoglobin_gr/dl (median[IQR]) | 13.6 (12.1, 14.9) | 3196 | 12.1 (10.3, 13.7) | 2855 |
| Hematocrit_% (median percentage [IQR]) | 40.2 (36.1, 44.0) | 3161 | 36.5 (31.3, 40.8) | 2850 |
| Platelets_ul (median[IQR]) | 228500.0 (176000.0, 297000.0) | 3230 | 283000.0 (202000.0, 378000.0) | 2890 |
| Prothrombin Time_seconds (median[IQR]) | 13.2 (12.0, 14.6) | 2540 | 13.2 (12.0, 15.1) | 1503 |
| Partial Thromboplastin Time_seconds (median[IQR]) | 31.8 (28.0, 36.8) | 2352 | 34.0 (29.0, 42.0) | 1310 |
| INR (median[IQR]) | 1.1 (1.0, 1.2) | 2408 | 1.1 (1.0, 1.3) | 1391 |
| Creatinine_mg/dl (median[IQR]) | 0.9 (0.7, 1.3) | 3193 | 0.9 (0.7, 1.3) | 2690 |
| Serum Sodium_mmol/l (median[IQR]) | 137.0 (134.0, 140.0) | 2910 | 138.0 (136.0, 141.0) | 2494 |
| Serum Potassium_mmol/l (median[IQR]) | 4.1 (3.8, 4.6) | 2906 | 4.2 (3.8, 4.7) | 2509 |
| Blood urea nitrogen_mg/dl (median[IQR]) | 18.0 (12.5, 28.9) | 2990 | 21.3 (14.3, 38.5) | 2506 |
| Lactic dehydrogenase_UI/l (median[IQR]) | 369.0 (269.0, 515.2) | 2704 | 293.0 (220.0, 440.8) | 1806 |
| Aspartate Transaminase_u/l (median[IQR]) | 42.0 (29.0, 65.8) | 2659 | 36.1 (24.0, 61.0) | 1630 |
| Alanine Aminotransferase_ u/l (median[IQR]) | 37.0 (24.0, 60.0) | 2583 | 44.1 (27.0, 79.8) | 1543 |
| Glucose_Glycemia_mg/dl (median[IQR]) | 125.0 (103.0, 175.0) | 2301 | 119.0 (95.7, 158.0) | 1671 |
| Arterial Blood Gas | | 3260 | | 3260 |
| 0. No | 676 (20.7) | | 1371 (42.1) | |
| 1. Yes | 2584 (79.3) | | 1889 (57.9) | |
| PH (median[IQR]) | 7.4 (7.4, 7.5) | 2519 | 7.4 (7.4, 7.4) | 1873 |
| Oxygen Blood Pressure_mmHg (median[IQR]) | 71.0 (58.7, 87.0) | 2576 | 74.0 (62.0, 88.0) | 1888 |
| Partial Pressure of Carbon Dioxide_mmHg (median[IQR]) | 32.3 (28.4, 37.0) | 2563 | 37.0 (33.0, 43.1) | 1886 |
| Bicarbonate_HCO3_mmol/L (median[IQR]) | 21.7 (19.2, 24.0) | 2477 | 23.5 (20.8, 26.2) | 1853 |
| PCT Test | | 3260 | | 3260 |
| 0. No | 455 (14.0) | | 1082 (33.2) | |
| 1. Sensitive PCR | 1739 (53.3) | | 1416 (43.4) | |
| 2. Ultrasensitive PCR | 1066 (32.7) | | 762 (23.4) | |
| Troponin Test | | 3260 | | 3260 |
| 0. No | 1174 (36.0) | | 2365 (72.5) | |
| 1. Troponin I | 376 (11.5) | | 99 (3.0) | |

| | | | | |
|---|---|---|---|---|
| 2. Troponin T | 143 (4.4) | | 54 (1.7) | |
| 3. Ultrasensitive Troponin I | 1216 (37.3) | | 489 (15.0) | |
| 4. Ultrasensitive Troponin T | 351 (10.8) | | 253 (7.8) | |
| Natriuretic Peptide Test | | 3260 | | 3260 |
| 0. No | 2711 (83.2) | | 3026 (92.8) | |
| 1. BNP | 94 (2.9) | | 50 (1.5) | |
| 2. NT-proBNP | 455 (14.0) | | 184 (5.6) | |
| CPK_U/L (median[IQR]) | 104.0 (53.0, 261.0) | 1281 | 77.0 (34.7, 256.5) | 683 |
| D-dimer_ug/mL (median[IQR]) | 0.8 (0.4, 1.5) | 2534 | 0.9 (0.4, 2.0) | 1663 |
| Fibrinogen_mg/dL (median[IQR]) | 550.0 (436.0, 663.0) | 1244 | 457.5 (354.0, 574.8) | 766 |
| Ferritin_ng/mL (median[IQR]) | 820.0 (396.0, 1481.5) | 2347 | 759.5 (367.2, 1333.0) | 1502 |
| Sensitive PCR_mg/dl (median[IQR]) | 10.1 (4.5, 20.0) | 1734 | 2.5 (0.8, 7.9) | 1415 |
| Ultrasensitive PCR_mg/dl (median[IQR]) | 11.6 (4.5, 22.2) | 1064 | 2.8 (0.8, 9.0) | 760 |
| Troponin I_ng/mL (median[IQR]) | 0.0 (0.0, 0.1) | 375 | 0.1 (0.0, 0.3) | 99 |
| Troponin T_ng/mL (median[IQR]) | 0.0 (0.0, 0.1) | 143 | 0.0 (0.0, 0.0) | 54 |
| Ultrasensitive Troponin I_ng/mL (median[IQR]) | 0.0 (0.0, 0.0) | 1216 | 0.0 (0.0, 0.1) | 489 |
| Ultrasensitive Troponin T_ng/mL (median[IQR]) | 0.0 (0.0, 0.0) | 351 | 0.0 (0.0, 0.1) | 253 |
| BNP_pg/mL (median[IQR]) | 100.2 (40.0, 432.2) | 94 | 80.2 (33.0, 219.9) | 50 |
| NT_proBNP_pg/mL (median[IQR]) | 429.0 (88.0, 2744.5) | 455 | 427.5 (131.5, 2954.2) | 184 |
